# Negative impact of COVID-19 associated health system shutdown on patients diagnosed with colorectal cancer: a retrospective study from a large tertiary center in Ontario, Canada

**DOI:** 10.1101/2021.07.30.21261400

**Authors:** Catherine L. Forse, Stephanie Petkiewicz, Iris Teo, Bibianna Purgina, Kristina-Ana Klaric, Tim Ramsay, Jason K. Wasserman

## Abstract

**Background:** In March 2020, a directive to halt all elective and non-urgent procedures was issued in Ontario, Canada because of COVID-19. The directive caused a temporary slowdown of screening programs including surveillance colonoscopies for colorectal cancer (CRC). Our goal was to determine if there was a difference in patient and tumour characteristics between CRC patients treated surgically prior to the COVID-19 directive compared to CRC patients treated after the slowdown.

**Methods:** CRC resections collected within the Champlain catchment area of eastern Ontario in the six months prior to COVID-19 (August 1, 2019-January 31, 2020) were compared to CRC resections collected in the six months post-COVID-19 slowdown (August 1, 2020-January 31, 2021). Clinical (e.g. gender, patient age, tumour site, clinical presentation) and pathological (tumour size, tumour stage, nodal stage, lymphovascular invasion) features were evaluated using chi square tests, T-tests and Mann-Whitney tests where appropriate.

**Results:** 343 CRC specimens were identified (175 pre-COVID-19, 168 post-COVID-19 slowdown). CRC patients treated surgically post-COVID-19 slowdown had larger tumours (44 mm vs. 35 mm; p = 0.0048) and were more likely to have presented emergently (24% vs .10%; p < 0.001). While there was a trend towards higher tumour stage, nodal stage, and clinical stage, these differences did not reach statistical significance. Other demographic and pathologic variables including patient gender, age, and tumour site were similar between the two cohorts.

**Interpretation:** The COVID-19 slowdown resulted in a shift in the severity of disease experienced by CRC patients in Ontario. Pandemic planning in the future should consider the long-term consequences to cancer diagnosis and management.

## Introduction

Colorectal cancer (CRC) is the third most common type of cancer in Canada and a leading cause of cancer death and morbidity worldwide.^1,2^ In many countries including Canada, the COVID-19 pandemic caused governments to limit access to non-essential health care services including colonoscopies necessary for CRC diagnosis. Unfortunately, this has resulted in a significant increase in the number of patients requiring screening, a backlog that will take months if not years to clear with the resources currently available.^3^ One study out of the United Kingdom reported a 72% decrease in the number of CRCs diagnosed at the beginning of the pandemic compared to the pre-pandemic period, a startling observation likely secondary to reduced screening.^4^

The number of colon cancers that will be missed due to delayed screening is difficult to estimate but predictions are dire. Models developed by several groups suggest that the cancellation of nond-urgent procedures and delayed screening will result in a significant increase in both the incidence of CRC and mortality once normal services resume.^5-7^ In addition, a greater percentage of patients are expected to present at a more advanced stage (stage III or IV).^5^

In March 2020, the Chief Medical Officer of Health for the Province of Ontario, Canada, ordered health care providers to stop performing all non-urgent and elective medical procedures (Directive #2). In the Champlain catchment area of eastern Ontario, the order allowed colonoscopies to continue for patients who had clinical features highly concerning for malignancy (e.g. rectal bleeding); however, screening colonoscopies for asymptomatic patients were immediately suspended until further notice. The purpose of this order was to ensure that hospitals had sufficient resources to manage and treat the large number of people who were expected to develop COVID-19 and to continue functioning in the face of unprecedented demand. However well intentioned, the order could also prevent people with other life-threatening but undiagnosed conditions such as cancer from accessing appropriate care in a timely manner.

In this study we sought to determine the impact of Directive #2 on patients diagnosed and surgically treated in eastern Ontario. Specifically, we compared the clinical and pathologic characteristics of patients treated prior to the cancellation of non-urgent and elective procedures in March 2020 to those treated after the resumption of normal services in August 2020.

## Methods

### Study population

All patients who underwent surgery for CRC within a region of eastern Ontario known as the Champlain local health integration network between August 1, 2019 and January 31, 2021 were included in this retrospective study. Patients were excluded if they were diagnosed and treated for CRC previously and therefore were already known to the health care system. The study was divided into three time periods. The “pre-slowdown” period included all patients treated surgically for CRC between August 1, 2019 and January 31, 2020. The pre-slowdown period preceded the start of the pandemic in Canada and the order to limit services. The “slowdown period” was defined as March 1, 2020 to July 31, 2020. The period includes the time covered by the original order and the time covered by a second order which allowed the slow restart of non-urgent and elective procedures. Because the restart allowed for only gradual resumption of services, the end of the slowdown period coincides with the time when hospital services were returning to pre-slowdown levels. The “post-slowdown” period included all patients diagnosed between August 1, 2020 and January 31, 2021. Most non-urgent and elective procedures including screening colonoscopies had resumed by the beginning of the post-slowdown period. Our analysis compared the characteristics of patients treated surgically in the pre-slowdown cohort and the post-slowdown cohort. Records for patients diagnosed during the slowdown period were retained for subsequent analysis.

### Data collection

All patient information (age, gender, clinical presentation, colonoscopy findings, neoadjuvant treatment) was extracted from The Ottawa Hospital electronic medical record database. All patients included in this study underwent surgical resection of the primary tumour either between August 1, 2019, and January 31, 2020 or August 1, 2020 and January 31, 2021. The surgical specimens were evaluated by pathologists at the Department of Pathology and Laboratory Medicine at the Ottawa Hospital. Information regarding surgical procedure, tumour location, tumour size, depth of invasion, lymphovascular invasion, lymph node metastases, and mesenteric tumour deposits was extracted from the final pathology report. Pathological tumour stage, nodal stage and clinical stage were assigned as per the AJCC 8^th^ edition TNM staging manual. Available imaging reports were used to determine the presence of distant metastases; however, this information was also occasionally found in the pathology report due to the presence of concurrent surgical specimens (e.g., liver resection).

An emergent presentation was defined as clinical and/or radiological evidence of severe bowel obstruction or bowel perforation requiring emergent, life-saving surgery. This information was obtained by reviewing both the clinical notes, colonoscopy reports and available abdominal imaging.

### Ethics approval

This study was approved by the Ottawa Health Science Network Research Ethics Board (20200331-01H).

### Statistical analysis

Categorical data are presented as number of patients and percentages. Continuous data are shown as either mean ± SD. Categorical and continuous variables were compared using the chi-square test and the Student’s t-test, respectively. The ordinal variables tumour stage, nodal stage, and clinical stage were compared pre-slowdown versus post-slowdown using Mann-Whitney tests. A value of p < 0.05 was considered statistically significant. Data were analysed using the Statistical Package for Social Sciences (SPSS) (IBM SPSS Statistics for Windows, version 25.0 (IBM Corp.).

## Results

A total of 343 patients were included in this study; 175 patients underwent CRC surgery during the pre-slowdown cohort and 168 patients underwent CRC surgery during the post-slowdown cohort. A summary of the data is presented in Table 1.

**Table.** Clinical and pathologic characteristics.

|  | Both cohorts | Pre-slowdown cohort | Post-slowdown cohort | p-value |
| --- | --- | --- | --- | --- |
| Number of patients | 343 (100) | 175 (51) | 168 (49) |  |
| Age, years (mean ± SD) | 68 ± 12 | 67±11 | 69±14 | 0.172 |
| Gender |  |  |  | 0.908 |
| Male | 193 (56) | 99 (57) | 94 (56) |  |
| Female | 150 (44) | 76 (43) | 74 (44) |  |
| Emergency presentation | 62 (18) | 17 (10) | 41 (24) | **< 0.001** |
| Perforation | 18 (5) | 8 (5) | 12 (7) | 0.310 |
| Obstruction | 38 (11) | 9 (5) | 29 (17) | **< 0.001** |
| Tumour location |  |  |  | 0.118 |
| Ascending colon | 120 (35) | 52 (30) | 68 (40) |  |
| Transverse colon | 21 (6) | 12 (7) | 9 (5) |  |
| Descending colon | 8 (2) | 6 (3) | 2 (1) |  |
| Sigmoid colon | 96 (28) | 48 (27) | 48 (29) |  |
| Rectum | 97 (28) | 57 (33) | 40 (24) |  |
| No neoadjuvant therapy | 267 (78) | 128 (73) | 139 (83) | **0.032** |
| Neoadjuvant chemotherapy | 65 (19) | 46 (26) | 19 (11) | **< 0.001** |
| Neoadjuvant radiation therapy | 57 (17) | 36 (21) | 21 (13) | **0.044** |
| Tumour size, mm (mean ± SD) | 39 ± 29 | 35 ± 28 | 44 ± 30 | **0.007** |
| Lymphovascular invasion | 155 (45) | 74 (42) | 81 (48) | 0.270 |
| Tumour deposits | 46 (13) | 24 (14) | 22 (13) | 0.866 |
| Nodal metastases | 120 (35) | 57 (33) | 63 (38) | 0.339 |
| Distant metastases | 33 (10) | 18 (10) | 15 (9) | 0.670 |
| Pathologic tumour stage |  |  |  | 0.160 |
| T0 | 18 (5) | 12 (7) | 6 (4) |  |
| Tis | 5 (1) | 2 (1) | 3 (2) |  |
| T1 | 44 (13) | 25 (14) | 19 (11) |  |
| T2 | 54 (16) | 28 (16) | 26 (15) |  |
| T3 | 144 (42) | 72 (41) | 72 (43) |  |
| T4a | 58 (17) | 28 (16) | 30 (18) |  |
| T4b | 19 (6) | 8 (5) | 11 (7) |  |
| Pathologic nodal stage |  |  |  | 0.630 |
| N0 | 177 (52) | 91 (52) | 86 (51) |  |
| N1a | 47 (14) | 21 (12) | 26 (15) |  |
| N1b | 29 (8) | 17 (10) | 12 (7) |  |
| N1c | 11 (3) | 8 (5) | 3 (2) |  |
| N2a | 22 (6) | 10 (6) | 12 (7) |  |
| N2b | 22 (6) | 9 (5) | 13 (8) |  |
| Stage |  |  |  | 0.380 |
| 0 | 17 (5) | 11 (6) | 6 (4) |  |
| I | 90 (26) | 50 (29) | 40 (24) |  |
| II | 106 (31) | 49 (28) | 57 (34) |  |
| III | 98 (29) | 47 (27) | 51 (30) |  |
| IV | 32 (9) | 18 (10) | 14 (8) |  |

### Clinical characteristics

The average patient age in this study was 68 ± 12 years. 193 of the patients (56%) were male and 150 of the patients (44%) were female. There was no significant difference between the post-slowdown cohort and the pre-slowdown cohort in terms of patient age or gender. A total of 62 patients (18%) had an emergent presentation; 18 patients (5%) with evidence of perforation and 38 patients (11%) with evidence of severe obstruction. Patients in the post-slowdown cohort were more likely to present emergently compared to patients in the pre-slowdown cohort (24% versus 10%; p < 0.001). This difference was driven primarily by the higher number of patients in the post-slowdown cohort who presented with severe bowel obstruction (17% versus 5%; p < 0.001). In contrast, there was no difference between the two cohorts in terms of the number of patients who presented with bowel perforation (7% versus 5%; p = 0.310).

Overall, the ascending colon was the most common tumour location (35%) followed by the sigmoid colon (28%) and rectum (28%). Patients in the post-slowdown cohort were more likely to have a tumour in the ascending colon (40% versus 30%) while patients in the pre-slowdown cohort were more likely to have a tumour in the rectum (24% versus 33%). These differences were not statistically significant (p = 0.118). Significantly fewer patients in the post-slowdown cohort received neoadjuvant chemotherapy (11% versus 26%, p <0.001) or neoadjuvant radiation therapy (13% versus 21%, p <0.044) compared to the patients in the pre-slowdown cohort.

### Pathologic characteristics

The average tumour size among all patients in this study was 39 ± 29 mm. Patients in the post-slowdown cohort had larger tumours compared to patients in the pre-slowdown cohort (44 mm versus 35 mm; p = 0.0048). Lymph node metastases were identified in 120 patients (35%) overall. There was no significant difference between the post-slowdown cohort and pre-slowdown cohort in the percentage of patients with either lymph node (38% versus 33%; p = 0.339) or distant metastases (9% versus 10%; p = 0.670). Lymphovascular invasion and tumour deposits also did not differ between the two cohorts.

Tumour stage and nodal stage were classified according to the American Joint Committee on Cancer 8^th^ edition. Overall, 18 (5%), 5 (1%), 44(13%), 54 (16%), 144 (42%), 58 (17%), and 19 (6%) tumours were classified as T0, Tis, T1, T2, T3, T4a, or T4b, respectively. Similarly, 177 (52%), 47 (14%), 29 (8%), 11 (3%), 22 (6%), and 22 (6%) cases were classified as N0, N1a, N1b, N1c, N2a, or N2b, respectively. When grouped according to clinical stage, 17 (5%), 90 (26%), 106 (31%), 98 (29%), and 32 (9%) patients were classified as stage 0, I, II, III, and IV, respectively. While there was a trend towards higher stage at presentation in the post-slowdown cohort, the difference was not statistically significant for tumour stage (Figure 1a; p = 0.16), nodal stage (Figure 1b; p = 0.38), or clinical stage (Figure 1c; p = 0.63).

**Figure.**
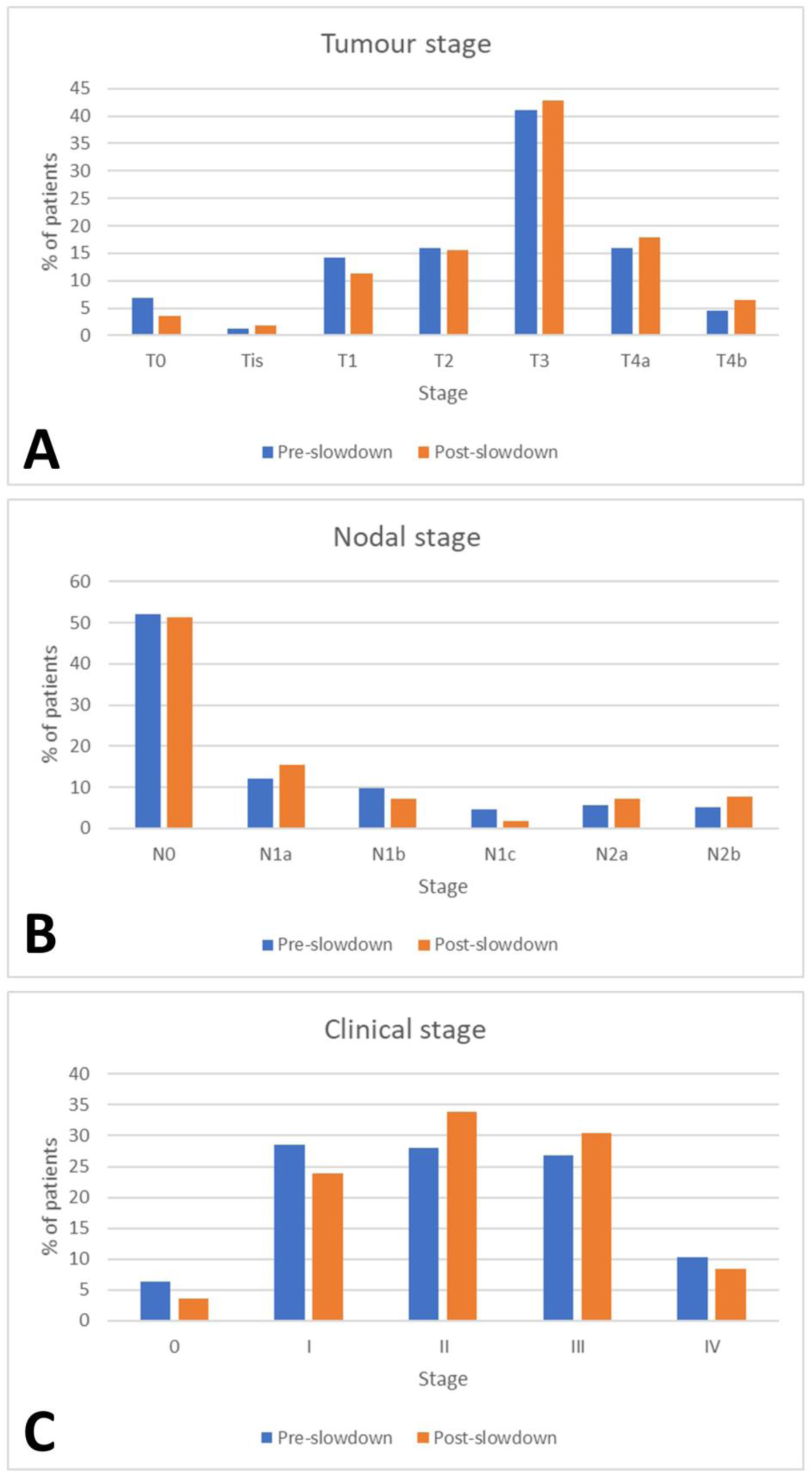
A comparison of tumour, nodal, and clinical stage.

## Discussion

Our study found that in patients surgically treated for CRC after cessation of non-urgent and elective procedures, a higher proportion presented emergently and had larger tumours than those who were treated prior to cessation of non-urgent and elective procedures. We also noted a trend towards higher tumour stage, nodal stage, and clinical stage. These results support the argument that government policies created in response to the COVID-19 pandemic to free up resources and minimize transmission may be inadvertently causing harm by delaying screening for patients with CRC.

The percentage of patients with CRC who present emergently varies from 9-32%, with one study estimating the proportion of CRC patients presenting emergently in Ontario as 18.7%.^8,9^ Even when successfully managed in the acute setting, patients who present emergently are more likely to die in the post-operative period compared to patients who present on an elective basis.^10,11^ With 10% of patients in the pre-slowdown cohort presenting emergently, our study is consistent with previously published studies from comparable populations. In contrast, the more than two-fold increase in the percentage of patients presenting emergently in the post-slowdown cohort places this group near the high end of the reported range and in line with populations studied prior to the introduction of large scale screening programs.^12^

The risk of obstruction or perforation varies based on the location of the tumours: hepatic flexure and descending colon tumours increase the risk for obstruction while cecal, descending colon, sigmoid colon tumours increase the risk for perforation. An increased risk of emergent presentation is also associated with more advanced tumours, female gender, and older patient age.^8,13^ None of these variables differed significantly between the patients in the pre-slowdown and post-slowdown cohorts in this study. Conversely, screening programs, such as those that were put on hold during the slowdown period, are associated with lower rates of emergent presentation as tumours and pre-cancerous lesions are identified and removed at an earlier stage.^12,14-16^

According to our data, patients in the post-slowdown cohort had tumours that were on average 25% larger than patients in the pre-slowdown cohort. The reduced number of patients receiving neoadjuvant therapy in the post-shutdown cohort may have marginally contributed to the differences in average tumour size between the two populations; however, the vast majority (78% of patients) did not receive neoadjuvant treatment.

Several studies have examined the growth of colon cancers over time and have found that the doubling time of a tumour is related to its stage. One retrospective radiographic study found a mean doubling time of 26.5 months for intramucosal adenocarcinoma, 25.9 months for carcinoma invasive into the submucosa, and 12.3 months for more advanced disease.^17^ Similarly, a meta analysis found that the doubling time ranged from 9.4 to 55.4 months for intramucosal adenocarcinoma and 4.7 to 12.2 months for advanced cancer.^17^ Thus, according to these reports, it is conceivable that an already advanced cancer could increase in size by 25% over a period of 6-months.

Although not statistically significant, it is interesting that more patients in the post-slowdown cohort presented with right-sided colon cancer than in the pre-slowdown cohort. Right-sided colon cancers are often diagnosed at a more advanced stage as they usually present with milder clinical symptoms (e.g. anemia, weight loss). Since colonoscopies during the slowdown were limited to patients who were symptomatic, it is logical that these patients would not have presented to the health care system until they had severe complications.

According to the TNM classification, the tumour stage reflects vertical tumour growth into the wall of the bowel. While tumour size is technically not a part of criteria used to determine the tumour stage, tumour size has been shown to correlate with vertical growth and larger tumours typically have invaded further into or beyond the wall at the time of diagnosis.^18^ A similar correlation has been shown for tumour size and nodal stage.^19^ While a visual inspection of our tumour stage, nodal stage, and clinical stage results appears to suggest a consistent pattern of higher stages in the post-slowdown period than the pre-slowdown period, none of these were statistically significant. Nonetheless, when combined with the larger tumour size and higher rate of emergent presentation post-slowdown, our results suggest that patients in the post-slowdown cohort had more advanced disease than their counterparts in the pre-slowdown cohort.

An important limitation of the current study is that, while we were able to show significant differences between the post-slowdown and pre-slowdown cohorts, we cannot identify a specific cause. One possible explanation is that the lack of screening procedures during the slowdown period resulted in more cancers going undetected at an earlier stage and that this led to a disproportionate number of advanced cancers in the post-slowdown cohort. This explanation would apply to those patients who would have normally undergone a screening procedure but were delayed/cancelled due to the slowdown. Another possible explanation is that some people may have been reluctant to seek medical attention. This may have been out of personal fear of COVID-19 or out of respect for government messaging to stay at home. Thus, some people may have discounted early warning signs and only presented to hospital once they had developed “alarm” symptoms (e.g., persistent abdominal pain). Unfortunately, both explanations involve system failures at a population level.

In summary, our findings demonstrate that health care policies put in place at the beginning of the COVID-19 pandemic in Ontario, Canada were ultimately associated with more advanced disease in patients diagnosed with CRC. Worsening cancer burden not only increases patient morbidity and mortality, but also has long-term social and economic effects on oncological care. Governments and health authorities should consider these results when creating emergency pandemic protocols in the future. In particular, efforts to continue cancer screening procedures with minimal interruption may help to minimize the long-term effects on cancer patient health.

## Data Availability

Data not available.

